# Mental Health Status and its Associated Factors among Women with Fertility Issues Visiting Fertility Center in Kathmandu Valley: A Cross-Sectional Study

**DOI:** 10.1101/2025.11.04.25339529

**Authors:** Pragya Kunwar, Amrita Ghimire, Aakriti Bharati, Amrit Bist, Laxmi Gautam

## Abstract

**Background:** Infertility poses a multidimensional burden, especially for women in low- and middle-income countries, where it is often associated with social stigma, emotional distress, and limited support. This study aimed to assess the mental health status and its associated factors among women with fertility issues visiting fertility center in the Kathmandu Valley.

**Methods:** A cross-sectional study was conducted from November 2023 to February 2024 among 143 women with fertility issues visiting a fertility center in the Kathmandu Valley. Nepali versions of the DASS-21 were administered to 143 women visiting the fertility center to collect data. Descriptive and inferential statistics were performed using SPSS version 22.0. The logistic regression model was applied to examine the factors associated with the outcome variable using an adjusted odds ratio with a 95% CI, and a p-value < 0.05 was considered statistically significant.

**Results:** A majority of respondents (82.5%) reported experiencing some level of anxiety, followed by 51.7% with stress and 35.0% with depression. Women who felt insecure about not having a child had significantly higher odds of stress (AOR=3.19), while those facing social stigmas also had elevated odds of stress (AOR=2.91) and anxiety (AOR=1.55). Experiencing abuse was strongly associated with stress (AOR=12.76). Women who had never conceived were more likely to experience anxiety (AOR=2.56), as were those facing financial burdens (AOR=4.78). Depression was more likely among women dissatisfied with their married life (AOR=4.30) and among those with a female factor of infertility (AOR=14.14) compared to unexplained infertility. Increased odds of depression were also associated with longer treatment duration (>5 years) (AOR=3.34) and higher treatment cost (>5 lakhs) (AOR=2.71).

**Conclusion:** Anxiety, stress and depression were found to be common among women with fertility issues. Higher psychological distress requires prompt actions to address the mental health needs of women who are battling with infertility.

## Background

Infertility has been recognized as a public health issue worldwide. According to the World Health Organization (WHO), infertility is defined as a disease of the male or female reproductive system defined by the failure to achieve a pregnancy after 12 months or more of regular unprotected sexual intercourse [1, 2]. It is estimated that approximately one in every six people of reproductive age worldwide may experience infertility during their lifetime [1]. It is believed that a significant proportion of couples, approximately 21% experience phases of infertility throughout their lives [3]. It is a multidimensional stressor with several kinds of emotional adjustments [4]. It is associated with different problems like dysfunction in sexual relationships, anxiety, depression and difficulties in marital life [5]. In the general population, major depression is two to three times as common among women in comparison to men [6, 7]. Depression is one of the most common negative emotions that is associated with infertility, in which the local social and cultural context may influence [6–8].

The WHO’s depression statistics over the world vary by sex, with the prevalence among females being 50% higher than males [9]. In other nations, the occurrence rate of depression ranges from 5-12% for men and 10-25% for women who are facing infertility [10]. Notably, major depression appears to be two to three times more prevalent in women compared to men [11]. The prevalence of depression among women with fertility issues as found in studies by Lok et al., Fatemeh et al., and Al-Homaidan was 33%, 40.8%, and 53.8%, respectively [12–14]. After 4 to 6 years of infertility, depression and anxiety are most typical. Severe depression was reported in individuals who had been infertile for 7 to 9 years; 40.8% of women with fertility issues had depression, and 86.8% had anxiety [13].

The study among women with fertility issues in the Tertiary Care Hospital of Karachi, Pakistan, indicated that infertile (primary infertile group) women had the highest percentage of depression (severe-58.2%), anxiety (severe-57.3%), and stress (severe-50.0%) [15]. A similar conclusion was reached in Bangladesh (54.5%) and Iran studies (58.0%), and a lower percentage was identified in studies in Sweden (14.8%), Saudi Arabia (21.8%), China (45.0%), and Nepal (40.0%) [16–19].

Infertility can have negative social and psychological implications for an individual, ranging from divorce to more subtle types of social stigma that lead to isolation and mental distress [8]. A study in Pakistan showed that more than 67.7% of participants had disputes in interpersonal relationships [20]. One of the most challenging parts that women with fertility issues express is dealing with feelings of jealousy and envy while learning of other women’s pregnancies or being in the presence of others who have infants [3]. Some women might hide their pain from healthcare experts because they are anxious, nervous or being criticized, or fear being labeled as insane [8]. According to reports, up to 13% of women develop passive thoughts of suicide following an unsuccessful In-Vitro Fertilization (IVF) effort [8].

The social repercussions for South Asian women like Nepal living in patriarchal cultures, where motherhood is often the sole way for women to advance in the family and community, are potentially far greater than for women in Western communities [21]. The woman may be held responsible for infertility regardless of whether the medical cause is with her or with her spouse [22]. The marriage relationship is frequently compromised, even leading to intimate partner violence [22]. Infertility has received little attention in Nepal because it is not a life-threatening condition, although it causes severe community health issues such as depression, anxiety, spousal violence, and social isolation [23]. Despite this, there has been no successful program created in Nepal to address the many reasons for infertility [23]. In reality, women with fertility issues face the constant threat of divorce or remaining in a polygamous family, which forces them to lower their quality of life and become victims of such difficulties [24].

This study assessed the psychological consequences of infertility in terms of the mental health status of women with fertility issues due to their treatment, family, and societal factors. This research can aid in the development of specific psychological interventions, which are now lacking in the national public healthcare routine, for women battling infertility in order to help them manage potential mental health problems while meeting their reproductive goals.

## Methods

### Study design, population and setting

A cross-sectional study was carried out from October 2023 to April 2024 to assess the mental health status and its associated factors among women with fertility issues attending fertility centers in the Kathmandu Valley. The Kathmandu Valley spans an area of 899 square kilometers and consists of three districts: Kathmandu (395 sq km), Lalitpur (385 sq km), and Bhaktapur (119 sq km) [25]. As per the 2021 national census, the valley is home to over three million people, making it the most densely populated region in Nepal [26]. Due to this population density and the concentration of infrastructure and services, Kathmandu Valley hosts the majority of specialized healthcare services in the country, including fertility care. Out of the 53 identified infertility care service providers in Nepal, 28 are located within the Kathmandu Valley, highlighting its role as a central hub for infertility treatment.

### Sample size and sampling process

The initial sample size was calculated to be 374 using Cochran’s formula, assuming a 95% confidence interval (CI), a 5% margin of error, and an estimated proportion (p) of 58%, based on a previous study conducted among infertile women attending a tertiary care hospital in Pakistan [15]. A 10% non-response rate was also accounted for in this calculation. However, only 200 eligible clients were identified from the selected Vatsalya Health Care’s patient records. Given this finite population, the final sample size was recalculated using the finite population correction formula:

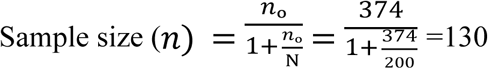

After adding 10% of non-response rate the final sample size for the study was determined to be 143.

The Kathmandu Valley was selected as the study area because fertility services in Nepal are largely concentrated in this region. Out of the 53 infertility service providers in the country, 28 are located within the valley. The Kathmandu Valley comprises three districts: Kathmandu, Lalitpur, and Bhaktapur. Among these districts, Kathmandu was selected as the study site due to its high concentration of fertility centers; 16 out of the 28 centers within the valley are located in Kathmandu district alone. Among these, Vatsalya Health Care Pvt. Ltd. (also known as Vatsalya Natural IVF Center) was chosen, as it had the highest patient flow among infertility centers in the district. A list of 200 eligible clients was obtained from the center, and 143 participants were then selected using simple random sampling. The ‘RANDBETWEEN’ function in Microsoft Excel was used to generate random numbers for participant selection.

### Data collection

A structured, validated Nepali version of the DASS-21 (N-DASS21) scale was used to collect the data for this study. DASS-21 is a psychological screening instrument capable of differentiating symptoms of DAS. Depression, anxiety, and stress are three subscales, and there are 7 items in each subscale. Each item is scored on a 4-point Likert scale, which ranges from 0 (did not apply to me at all) to 3 (applied to me very much) [27]. Scores for DAS were calculated by summing the scores for the relevant items and multiplying by two. A face-to-face interview with children was carried out by a trained public health professional. Pretesting of the questionnaire was done among 10% of the sample size among women with fertility issues visiting the Ishan Children Hospital and IVF center in Tokha, Kathmandu. DASS 21 is a valid standard scale frequently used by nurses, psychologists, and psychiatrists of Nepal [28]. The DASS-21 is a validated tool and has been used in different groups of the Nepalese population to identify symptoms of depression, anxiety and stress. Cronbach’s alpha coefficient for DASS 21 has been found to range from 0.76 to 0.91 [28–30].(S1 Text)

### Scoring and Interpretation

#### Depression

Dysphoria, hopelessness, devaluation of life, self-deprecation, lack of interest/involvement, anhedonia and inertia (Items 3, 5, 10, 13, 16, 17, 21).

#### Anxiety

Autonomic arousal, skeletal muscle effects, situational anxiety, and subjective experience of anxious affect. (Items 2, 4, 7, 9, 15, 19, 20).

#### Stress

Levels of chronic nonspecific arousal, difficulty relaxing, nervous arousal, and being easily upset/agitated, irritable/over-reactive and impatient (Items 1, 6, 8, 11, 12, 14, 18).

### Data management and analysis

A comprehensive codebook was developed prior to data entry to ensure consistency and standardization of the collected information. The data were initially entered into Microsoft Excel 2019, and once the dataset was cleaned and validated, it was imported into the Statistical Package for Social Sciences (SPSS) version 22.0 for statistical analysis. Descriptive statistics, including frequencies and percentages, were computed for all relevant dependent and independent variables. Bivariate analyses were conducted to assess associations between the outcome variable and each explanatory variable. Variables with p-value <0.25 in bivariate analysis were included in a multivariable binary logistic regression model to assess their independent effects on the outcome, with results presented as adjusted odds ratios (aORs) and 95% confidence intervals.(S1 Data)

To address potential multicollinearity among independent variables included in the multivariable model, the Variance Inflation Factor (VIF) was calculated. A threshold of VIF >2 was considered indicative of multicollinearity [31], and no multicollinearity was found between the variables. Variables with p-values <0.05 were taken as statistically significant.

## Results

The majority of the respondents were aged between 30-40 years (51.7%), followed by those aged 20-30 years (25.9%). In terms of ethnicity, the largest proportion belonged to the Janajati group (33.6%), followed by Chhetri (28.0%) and Brahmin (23.1%). A vast majority of the participants were Hindu (84.6%). In terms of educational attainment, secondary education was most common (41.3%), followed by a university degree (32.9%), a postgraduate degree (12.6%) and majority (56.6%) of participants lived in a joint family. The median age at marriage was 24 years (IQR ±5). More than half (55.9%) were married at the age of 24 or older, and 44.1% were married before age 24 years. Regarding the duration of marriage, one-third (33.6%) had been married for less than 5 years whereas almost one out of four (23.8%) were married for 15 years or more. In terms of permanent residence, 62.9% were from outside the Kathmandu Valley, 34.3% resided within the valley, and a small proportion (2.8%) reported their permanent address as being outside Nepal (Table 2).

**Table 1:** The Recommended Cut-off Scores for Conventional Severity Levels.

| Severity Level | Depression | Anxiety | Stress |
| --- | --- | --- | --- |
| Normal | 0-9 | 0-7 | 0-14 |
| Mild | 10-13 | 8-9 | 15-18 |
| Moderate | 14-20 | 10-14 | 19-25 |
| Severe | 21-27 | 15-19 | 26-33 |
| Extremely Severe | 28+ | 20+ | 34+ |

Source: Lovibond, S. H., & Lovibond, PF. Manual for the depression anxiety stress scales [29].

**Table 2:** Socio-demographic characteristics of the respondents.

| Variables | Frequency (n=143) | Percentage |
| --- | --- | --- |
| **Age in years (Min-23, Max-58)** |  |  |
| 20-30 | 37 | 25.9 |
| 30-40 | 74 | 51.7 |
| 40+ | 31 | 22.28 |
| **Ethnicity** |  |  |
| Brahmin | 33 | 23.1 |
| Chhetri | 40 | 28.0 |
| Madhesi | 12 | 8.4 |
| Dalit | 8 | 5.6 |
| Janajati | 48 | 33.6 |
| Others | 2 | 1.4 |
| **Religion** |  |  |
| Hindu | 121 | 84.6 |
| Buddhism | 11 | 7.7 |
| Christianity | 9 | 6.3 |
| Muslim and other | 2 | 1.4 |
| **Highest level of education** |  |  |
| No education (never been to school) | 1 | 0.7 |
| Basic Education | 18 | 12.6 |
| Secondary Education | 59 | 41.3 |
| University degree | 47 | 32.9 |
| Post graduate degree | 18 | 12.6 |
| **Family type** |  |  |
| Nuclear family | 62 | 43.4 |
| Joint family | 81 | 56.6 |
| **Age at marriage (in years)** |  |  |
| **(Median ± IQR: 24±5)** |  |  |
| <24 | 63 | 44.1 |
| ≥24 | 80 | 55.9 |
| **Duration of marriage (in years)** |  |  |
| <5 | 48 | 33.6 |
| 5-9.99 | 40 | 28.0 |
| 10-14.99 | 21 | 14.7 |
| ≥15 | 34 | 23.8 |
| **Permanent Address** |  |  |
| Inside valley | 49 | 34.3 |
| Outside valley | 90 | 62.9 |
| Out of Nepal | 4 | 2.8 |

Among the 143 women interviewed, almost half were (44.8%) working. Among the women working (n=64), the highest percentage (70.3%) were involved in Government/private service. More than one-third of the respondents were unemployed (38.5%), and a higher percentage of women (16.8%) had left work only due to treatment. Those who were working (n=64), almost half of respondents (45.3%), find it difficult to visit the clinic due to work, and more than half (56.5%) took daily leave from work to get treatment. The average monthly family income was NRS 60,000 ± 50,000, where more than half of respondents (54.5%) had a family income more than NRS 60,000 (Table 3).

**Table 3:**
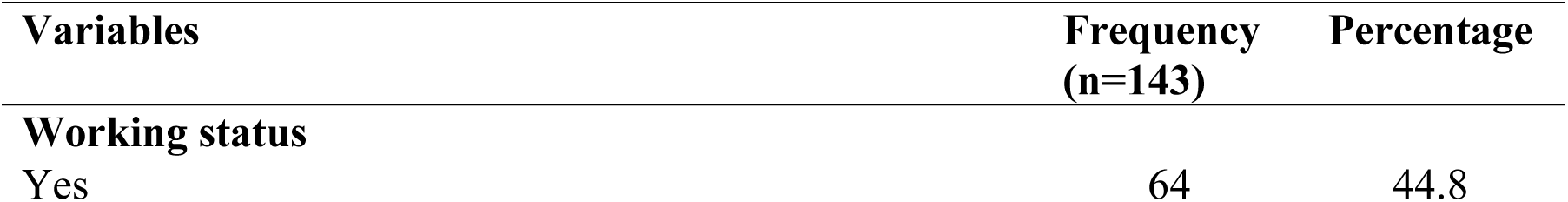
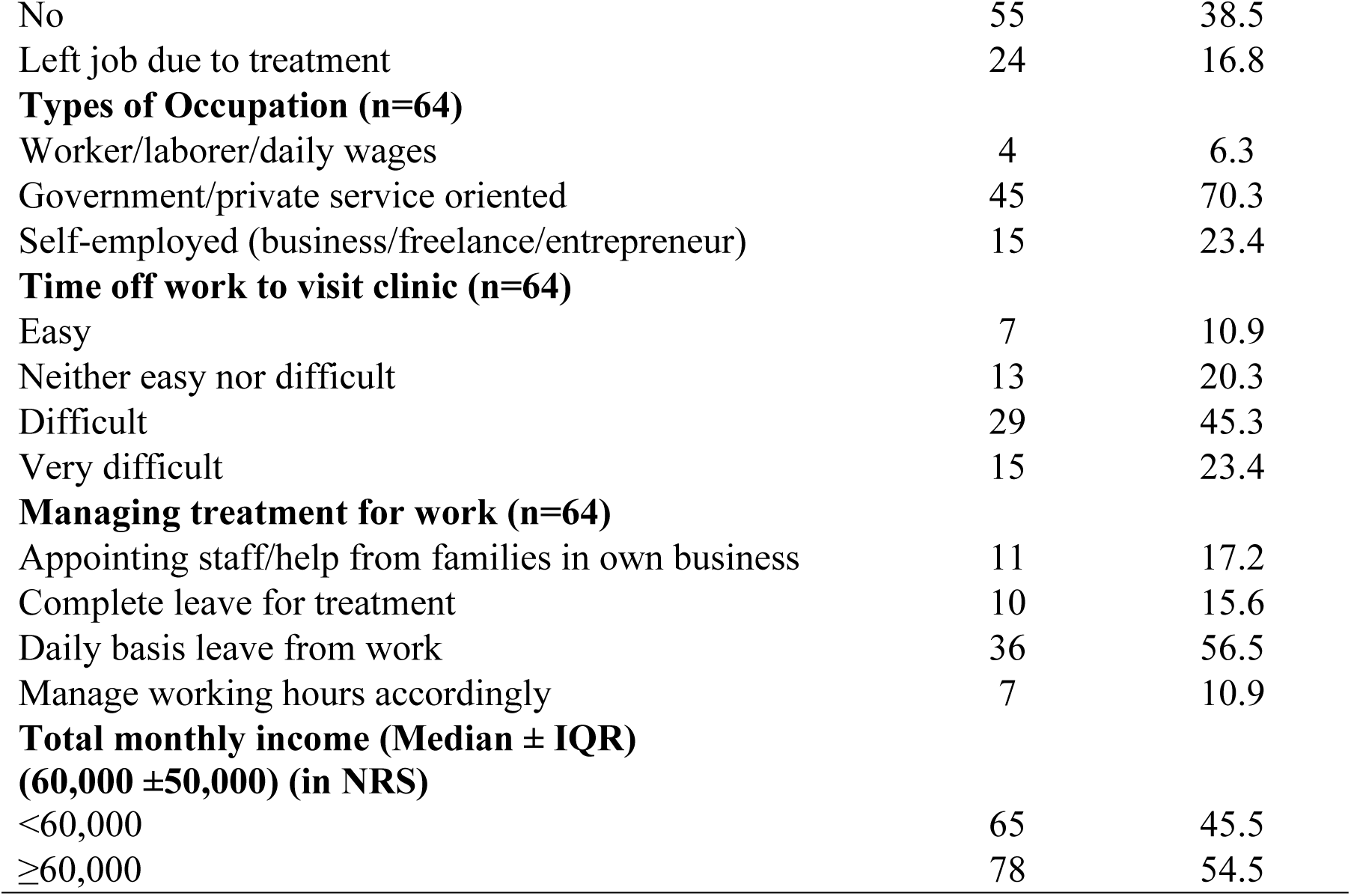
Socio-economic characteristics of the respondents.

| Variables | Frequency<br>(n=143) | Percentage |
| --- | --- | --- |
| **Working status** |  |  |
| Yes | 64 | 44.8 |
| No | 55 | 38.5 |
| Left job due to treatment | 24 | 16.8 |
| **Types of Occupation (n=64)** |  |  |
| Worker/laborer/daily wages | 4 | 6.3 |
| Government/private service oriented | 45 | 70.3 |
| Self-employed (business/freelance/entrepreneur) | 15 | 23.4 |
| **Time off work to visit clinic (n=64)** |  |  |
| Easy | 7 | 10.9 |
| Neither easy nor difficult | 13 | 20.3 |
| Difficult | 29 | 45.3 |
| Very difficult | 15 | 23.4 |
| **Managing treatment for work (n=64)** |  |  |
| Appointing staff/help from families in own business | 11 | 17.2 |
| Complete leave for treatment | 10 | 15.6 |
| Daily basis leave from work | 36 | 56.5 |
| Manage working hours accordingly | 7 | 10.9 |
| **Total monthly income (Median ± IQR)**<br>**(60,000 ± 50,000) (in NRS)** |  |  |
| <60,000 | 65 | 45.5 |
| ≥60,000 | 78 | 54.5 |

The majority of the respondents (77.6%) were satisfied with their marital life. Among unsatisfied women (n=7), 28.6% each said their husband’s habit of drinking alcohol and not having a child were their reasons. Having child was essential to most (86.7%) of the women. More than half of the respondents (58.7%) had never conceived in their lifetime, and the majority of respondents (72%) were insecure about not having a child. Above the three-fourth of the respondents (76.2%) had not noticed any changes in family’s behavior after the fertility issues. Among the respondents (n=34) who had faced changes, 41.18% were accused of not having an heir of the family, and 17.7% each were accused of being ill and faced different arguments, respectively. Whereas 14.7% of respondents’ families asked their husbands to remarry to have a child, and the remaining 8.9% had lost their importance and support from family as in earlier days (Table 4).

**Table 4:** Social characteristics of the respondents.

| **Variables** | **Frequency**<br>**(n=143)** | **Percentage** |
| --- | --- | --- |
| **Marital Satisfaction** |  |  |
| Yes | 111 | 77.6 |
| No | 7 | 4.9 |
| Neutral | 25 | 17.5 |
| **Reasons for dissatisfaction (n=7)** |  |  |
| Divorce and lost child in accident | 1 | 14.3 |
| No child | 2 | 28.6 |
| No support from family | 1 | 14.3 |
| Husband drinks a lot | 2 | 28.6 |
| Violence from husband's family | 1 | 14.3 |
| **Parity** |  |  |
| Never Conceived | 84 | 58.7 |
| Conceived Earlier but no live children | 45 | 31.5 |
| Had live children earlier but difficulty in conceiving now | 13 | 9.1 |
| Others (Adoption) | 1 | 0.7 |
| **Importance to have a child** |  |  |
| Very Important | 124 | 86.7 |
| Important | 19 | 13.3 |
| **Insecure about not having a child** |  |  |
| Yes | 103 | 72.0 |
| No | 40 | 28.0 |
| **Change in families behaviors** |  |  |
| Yes | 34 | 23.8 |
| No | 109 | 76.2 |
| **Types of changes (n=34)** |  |  |
| Accused of not having the heir of the family | 14 | 41.2 |
| Accused of being ill | 6 | 17.7 |
| Asked husband to remarry | 5 | 14.7 |
| Often arguments and no support | 6 | 17.7 |
| Not giving much importance as earlier | 3 | 8.8 |

More than one third of respondents (40.6%) families were neutral about their treatment for infertility. Half of the respondents (50.3%) had faced social stigmas during their lifetime due to fertility issues. Among them, 32.4% of women were judged on their personality and accused by negative comments, followed by 21.1% of women accused of being ‘*Bajh*’ and also 8.5% of respondents’ husbands were asked to remarry by the relatives and society.

Similarly, the majority of the respondents (70.6%) were involved in social activities. Among the respondents who were not involved, 28.6% of women had fear of judgments and stress from society. Well above three-fourth of the respondents (78.3%) felt comfortable sharing their personal feelings with anyone. Regarding the abuse faced due to fertility issue during their lifetime, 13.3% (n=19) of the women had positive response. Among which, reported abusers were majority society people (84.2%), followed by relatives (47.4%), in-laws and other family members as (26.3%). Among total self-reported abuse (n=19), around three-fourths of respondents (73.7%) faced both verbal and emotional abuse, 15.7% verbal abuse only (Table 5).

**Table 5:** Social characteristics of the respondents.

| **Variables** | **Frequency**<br>**(n=143)** | **Percentage** |
| --- | --- | --- |
| **Support of family (in-laws) for the treatment** |  |  |
| Not supportive at all | 2 | 1.4 |
| Not so supportive | 6 | 4.2 |
| Neither supportive nor unsupportive | 58 | 40.6 |
| Supportive | 21 | 14.7 |
| Very supportive | 56 | 39.2 |
| **Social stigmas faced due to infertility** |  |  |
| Yes | 71 | 50.3 |
| No | 72 | 49.7 |
| **Types of Social stigmas faced (n=71)** |  |  |
| Accused of being career-oriented and married late | 9 | 12.7 |
| Accused of being incomplete, wrong or 'papi' | 6 | 8.5 |
| Accused of being 'bajh' | 15 | 21.1 |
| Judgments on personality and negative comments | 23 | 32.4 |
| Second marriage suggestions to husband | 6 | 8.5 |
| Still not conceived after a long duration of marriage | 8 | 11.3 |
| Stereotypical judgments on IVF | 4 | 5.7 |
| **Involved in social activities** |  |  |
| Yes | 101 | 70.6 |
| No | 42 | 29.4 |
| **Reasons for not being involved in social activities (n=42)** |  |  |
| Fear of judgments and stress | 12 | 28.6 |
| Always asks for child | 4 | 9.5 |
| Jealous of other women and kids | 2 | 4.8 |
| No free time from household chores and office | 16 | 38.1 |
| Not interested in even participating | 8 | 19.1 |
| **Share feelings with anyone** |  |  |
| Yes | 112 | 78.3 |
| No | 31 | 21.7 |
| **Reasons for not sharing feelings with anyone (n=31)** |  |  |
| Not comfortable to share personal stuffs | 19 | 61.3 |
| Fear of judgments from others | 8 | 25.8 |
| Male factor infertility cases stereotypes | 4 | 12.9 |
| **Abuse due to infertility issues** |  |  |
| Yes | 19 | 13.3 |
| No | 124 | 86.7 |
| **Reported abusers\* (n=19)** |  |  |
| Husband | 1 | 5.26 |
| In-laws | 5 | 26.23 |
| Husband and other family members | 5 | 26.23 |
| Society | 16 | 84.21 |
| relatives | 9 | 47.37 |
| Others |  |  |
| **Types of abuses (n=19)** |  |  |
| Verbal | 3 | 15.7 |
| Emotional | 1 | 5.3 |
| Verbal and emotional | 14 | 73.7 |
| Sometimes physical | 1 | 5.3 |

Among 143 respondents, the majority of the respondents (90.2%) had primary infertility issues. More than one-fifth the respondents (42.7%) had fertility issues for more than 5 years. Among which the female infertility factor was higher percentage (62.9%) whereas 10.5% had the male factor. Half of the respondents (50.3%) had also undergone treatment before the specific period of time, with almost one-third (31.9%) got treatment already for 2 times. More than half of the respondents (51.7%) had IVF from couple’s own egg and sperm and two-fifth (40.6%) of them were seeking treatment for less than 2 years.

More than half of the respondents (56.6%) shared the infertility treatment procedure with family members. Among 62 respondents who hadn’t shared it with family, two-fifth of them (40.3%) had fear of judgments and unacceptance of infertility treatment due to different stereotypes in rural areas. More than half of the respondents (56.6%) had treatment cost less than NRS 5 lakhs, and nearly half of the respondents (49.7%) had a financial burden. Among which, one-third of respondents (66.2%) had a delay of treatment due to financial burden due to the treatment cost and more than one-third had plans for selling more land and properties for future treatment costs (Table 6).

**Table 6:** Treatment related characteristics of the respondents.

| **Variables** | **Frequency**<br>**(n=143)** | **Percentage** |
| --- | --- | --- |
| **Type of infertility** |  |  |
| Primary | 129 | 90.2 |
| Secondary | 14 | 9.8 |
| **Duration of infertility** |  |  |
| < 2 years | 50 | 35 |
| 2-5 years | 32 | 22.4 |
| > 5 years | 61 | 42.7 |
| **Factor of infertility** |  |  |
| Male | 15 | 10.5 |
| Female | 90 | 62.9 |
| Unexplained | 38 | 26.6 |
| **Treatment procedure (Recent)** |  |  |
| Timed intercourse/Ovulation induction | 1 | 0.7 |
| Intrauterine insemination (husband sperm) | 8 | 5.6 |
| Intrauterine insemination (Donor sperm) | 3 | 2.1 |
| In-vitro fertilization (Couple) | 74 | 51.7 |
| In-vitro fertilization (Donor sperm) | 10 | 7 |
| In-vitro fertilization (egg donation) | 45 | 31.5 |
| Intra-cytoplasmic sperm injection (husband sperm) | 1 | 0.7 |
| Intra-cytoplasmic sperm injection (donor sperm) | 1 | 0.7 |
| **Any treatment before** |  |  |
| Yes | 72 | 50.3 |
| No | 71 | 49.7 |
| **Treatment times (before) n=72** |  |  |
| once | 22 | 30.6 |
| 2 times | 23 | 31.9 |
| 3 times | 15 | 20.8 |
| 4 times | 5 | 6.9 |
| 5 times | 6 | 8.3 |
| 6 times | 1 | 1.4 |
| **Duration of getting treatment (all)** |  |  |
| < 2 years | 85 | 59.4 |
| 2-5 years | 26 | 18.2 |
| more than 5years | 32 | 22.4 |
| **Infertility shared with family** |  |  |
| Yes | 81 | 56.6 |
| No | 62 | 43.4 |
| **Reasons for not sharing with family (n=62)** |  |  |
| Fear of in-laws and other family members | 23 | 37.1 |
| Fear of judgments and unacceptance due to stereotypical thoughts in rural areas | 25 | 40.3 |
| Fear of treatment and its negative results | 9 | 14.5 |
| Secondary infertility | 3 | 4.8 |
| Male infertility factor | 2 | 3.2 |
| **Thought of sharing to family in the future (n=62)** |  |  |
| Yes | 2 | 3.2 |
| No | 60 | 96.8 |
| **Treatment cost (till now)** |  |  |
| <500,000 NRS | 81 | 56.6 |
| ≥500,000 NRS | 62 | 43.4 |
| **Financial Burden** |  |  |
| Yes | 71 | 49.7 |
| No | 72 | 50.3 |
| **Delay in treatment (n=71)** |  |  |
| Yes | 47 | 66.2 |
| No | 24 | 33.8 |
| **Managing the cost (n=71)** |  |  |
| All savings, salary and house rents | 31 | 43.7 |
| Borrowing from friends, closed ones | 8 | 11.3 |
| Complete loans | 20 | 28.2 |
| Selling properties, gold | 12 | 16.9 |
| **Future financial treatment plans (n=71)** |  |  |
| Savings and salaries | 12 | 16.9 |
| Loans to be added | 18 | 25.4 |
| Selling properties, lands | 27 | 38.02 |
| Business, rents from house | 9 | 12.7 |
| Ask to families | 5 | 7.0 |

Among the 143 respondents, 14% experienced severe stress, and 1.4% reported extremely severe stress. Regarding anxiety, 44.1% of respondents had moderate anxiety, followed by 14.7% with severe and 11.2% with extremely severe anxiety. In terms of depression, 19.6% of respondents reported mild depression, while 2.8% and 0.7% experienced severe and extremely severe depression, respectively. Overall, the majority of respondents (82.5%) exhibited some level of anxiety, followed by 51.7% with stress and 35.0% with some degree of depression (Table 7).

**Table 7:** DASS Tool Scale Analysis.

| Variables | Frequency (n=143) | Percentage |
| --- | --- | --- |
| **Stress Level** |  |  |
| Normal | 69 | 48.3 |
| Mild | 31 | 21.7 |
| Moderate | 21 | 14.7 |
| Severe | 20 | 14 |
| Extremely Severe | 2 | 1.4 |
| **Anxiety level** |  |  |
| Normal | 25 | 17.5 |
| Mild | 18 | 12.6 |
| Moderate | 63 | 44.1 |
| Severe | 21 | 14.7 |
| Extremely Severe | 16 | 11.2 |
| **Depression level** |  |  |
| Normal | 93 | 65 |
| Mild | 28 | 19.6 |
| Moderate | 17 | 11.9 |
| Severe | 4 | 2.8 |
| Extremely Severe | 1 | 0.7 |

The odd of having stress among women who were insecure about not having a child was 3.19 times (AOR=3.19, 95% CI=1.98-10.39) than those who weren’t insecure. Likewise, the odd of having stress among women facing social stigmas was 2.91 times (AOR=2.91, 95% CI= 0.93-9.09) as compared to the women not facing the social stigmas. Similarly, the odd of having stress among women facing abuse was 12.76 times higher (AOR=12.76, 95% CI=1.38-118.13) than women not facing the abuse during their lifetime, adjusting all other independent variables (Table 8).

**Table 8:**
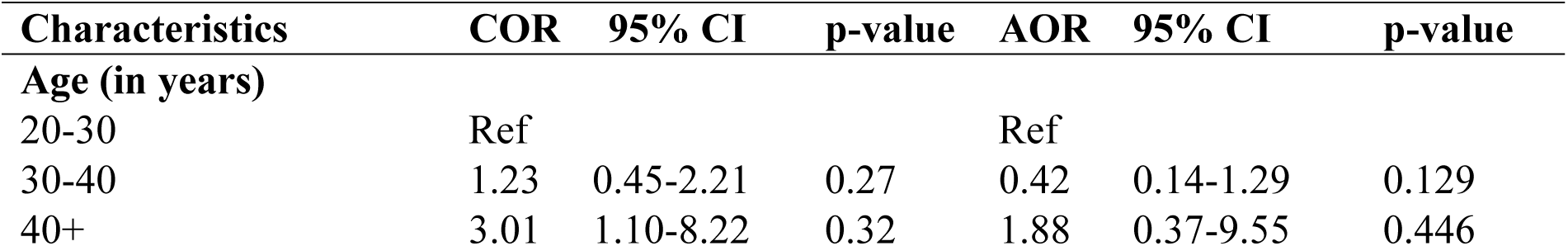
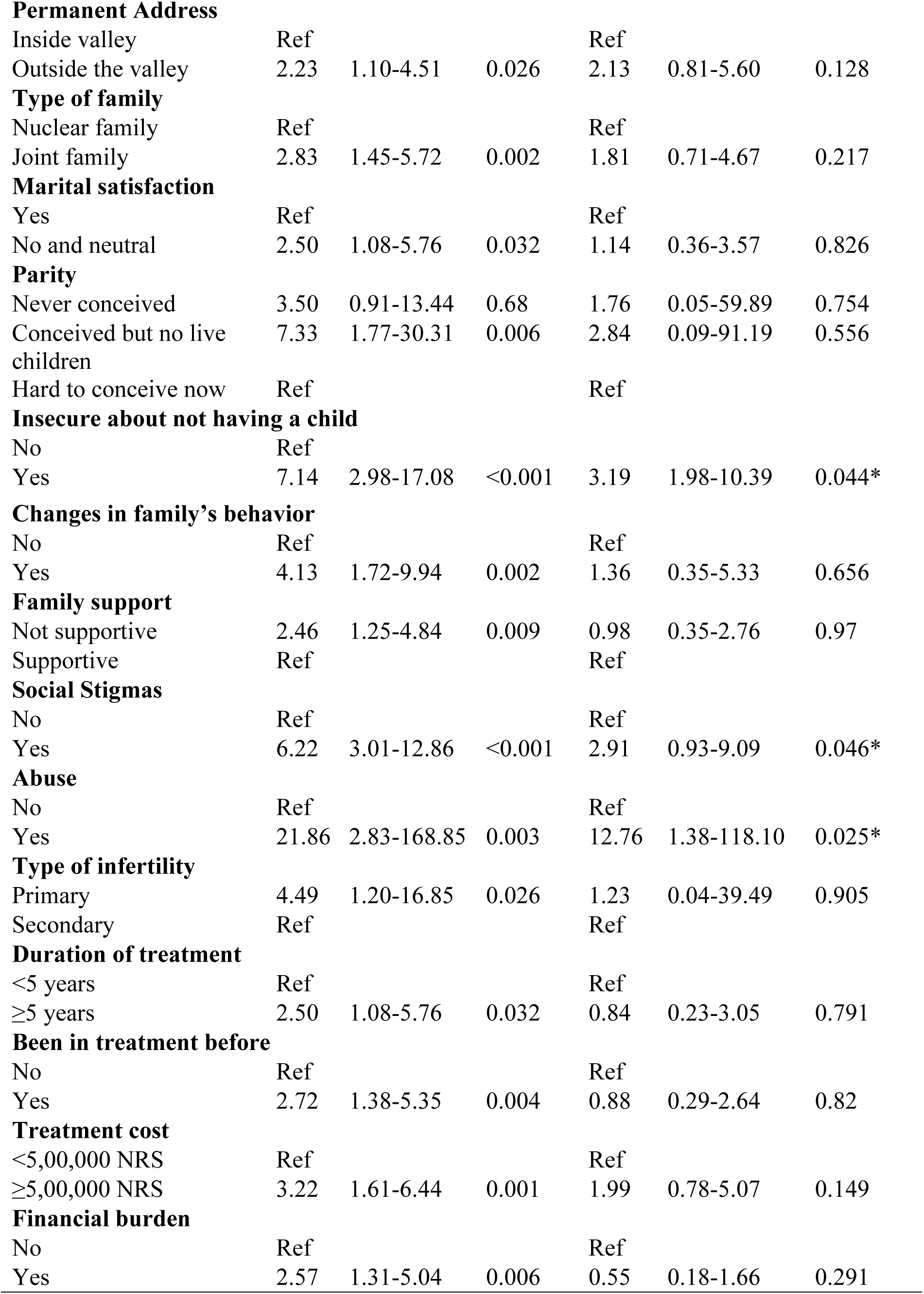
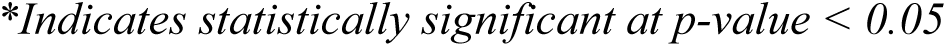
Multiple Logistics Regression between Stress and associated factors.

| Characteristics | COR | 95% CI | p-value | AOR | 95% CI | p-value |
| --- | --- | --- | --- | --- | --- | --- |
| **Age (in years)** |  |  |  |  |  |  |
| 20-30 | Ref |  |  | Ref |  |  |
| 30-40 | 1.23 | 0.45-2.21 | 0.27 | 0.42 | 0.14-1.29 | 0.129 |
| 40+ | 3.01 | 1.10-8.22 | 0.32 | 1.88 | 0.37-9.55 | 0.446 |
| **Permanent Address** |  |  |  |  |  |  |
| Inside valley | Ref |  |  | Ref |  |  |
| Outside the valley | 2.23 | 1.10-4.51 | 0.026 | 2.13 | 0.81-5.60 | 0.128 |
| **Type of family** |  |  |  |  |  |  |
| Nuclear family | Ref |  |  | Ref |  |  |
| Joint family | 2.83 | 1.45-5.72 | 0.002 | 1.81 | 0.71-4.67 | 0.217 |
| **Marital satisfaction** |  |  |  |  |  |  |
| Yes | Ref |  |  | Ref |  |  |
| No and neutral | 2.50 | 1.08-5.76 | 0.032 | 1.14 | 0.36-3.57 | 0.826 |
| **Parity** |  |  |  |  |  |  |
| Never conceived | 3.50 | 0.91-13.44 | 0.68 | 1.76 | 0.05-59.89 | 0.754 |
| Conceived but no live children | 7.33 | 1.77-30.31 | 0.006 | 2.84 | 0.09-91.19 | 0.556 |
| Hard to conceive now | Ref |  |  | Ref |  |  |
| **Insecure about not having a child** |  |  |  |  |  |  |
| No | Ref |  |  |  |  |  |
| Yes | 7.14 | 2.98-17.08 | <0.001 | 3.19 | 1.98-10.39 | 0.044* |
| **Changes in family's behavior** |  |  |  |  |  |  |
| No | Ref |  |  | Ref |  |  |
| Yes | 4.13 | 1.72-9.94 | 0.002 | 1.36 | 0.35-5.33 | 0.656 |
| **Family support** |  |  |  |  |  |  |
| Not supportive | 2.46 | 1.25-4.84 | 0.009 | 0.98 | 0.35-2.76 | 0.97 |
| Supportive | Ref |  |  | Ref |  |  |
| **Social Stigmas** |  |  |  |  |  |  |
| No | Ref |  |  | Ref |  |  |
| Yes | 6.22 | 3.01-12.86 | <0.001 | 2.91 | 0.93-9.09 | 0.046* |
| **Abuse** |  |  |  |  |  |  |
| No | Ref |  |  | Ref |  |  |
| Yes | 21.86 | 2.83-168.85 | 0.003 | 12.76 | 1.38-118.10 | 0.025* |
| **Type of infertility** |  |  |  |  |  |  |
| Primary | 4.49 | 1.20-16.85 | 0.026 | 1.23 | 0.04-39.49 | 0.905 |
| Secondary | Ref |  |  | Ref |  |  |
| **Duration of treatment** |  |  |  |  |  |  |
| <5 years | Ref |  |  | Ref |  |  |
| ≥5 years | 2.50 | 1.08-5.76 | 0.032 | 0.84 | 0.23-3.05 | 0.791 |
| **Been in treatment before** |  |  |  |  |  |  |
| No | Ref |  |  | Ref |  |  |
| Yes | 2.72 | 1.38-5.35 | 0.004 | 0.88 | 0.29-2.64 | 0.82 |
| **Treatment cost** |  |  |  |  |  |  |
| <5,00,000 NRS | Ref |  |  | Ref |  |  |
| ≥5,00,000 NRS | 3.22 | 1.61-6.44 | 0.001 | 1.99 | 0.78-5.07 | 0.149 |
| **Financial burden** |  |  |  |  |  |  |
| No | Ref |  |  | Ref |  |  |
| Yes | 2.57 | 1.31-5.04 | 0.006 | 0.55 | 0.18-1.66 | 0.291 |

*\*Indicates statistically significant at p-value < 0.05*

The odds of having anxiety among women who had never conceived were 2.56 times (AOR=2.56, 95% CI=1.65-36.09) whereas having conceived earlier but not having living children was 4.86 times (AOR=4.86, 95% CI= 2.61-90.76) than women that were having issues in conceiving in the present context. Likewise, the odds of having anxiety among women facing social stigmas were 1.55 times (AOR=1.55, 95% CI= 0.39-6.17) higher than women not facing the social stigmas. Similarly, the odd of having anxiety among women facing financial burden was 4.78 times (AOR=4.78, 95% CI= 1.05-21.89) higher than women not facing financial burden, adjusting for all other variables (Table 9).

**Table 9:**
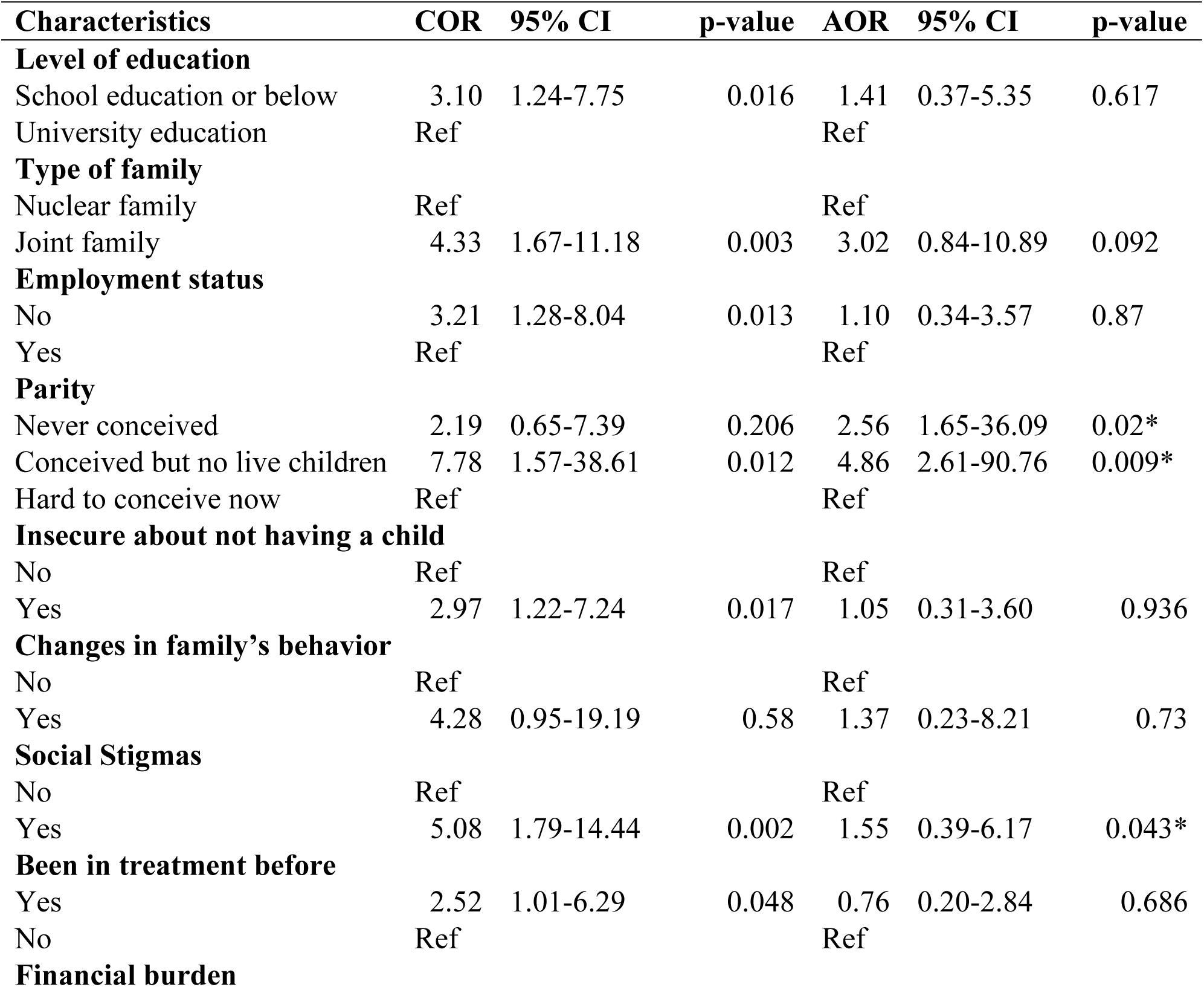
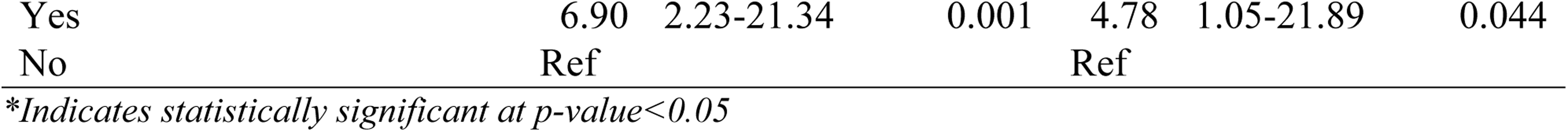
Multiple Logistic Regression between Anxiety and associated factors.

The odds of having depression among non-Hindu women were 7.61 times (AOR=7.61, 95% CI= 1.73-33.49) those of Hindu women. The odd of having depression among women who were not satisfied with their married life was 4.30 times (AOR=4.30, 95% CI= 1.12-16.50) that of satisfied women. Similarly, the odds of having depression among women having female factors of infertility were 14.14 times (AOR=14.14, 95% CI= 1.57-127.40) higher than for those with an unexplained factor of infertility. The odds of having depression among women having treatment duration more than 5 years were 3.34 times (AOR=3.34, 95% CI= 0.82-13.61) higher than women having treatment duration less than this. Likewise, the odd of having depression among women who had treatment costing more than five lakhs was 2.71 times (AOR=2.71, 95% CI= 0.21-26.74) higher than women having treatment costing less than that, adjusting other variables (Table 10).

**Table 10:**
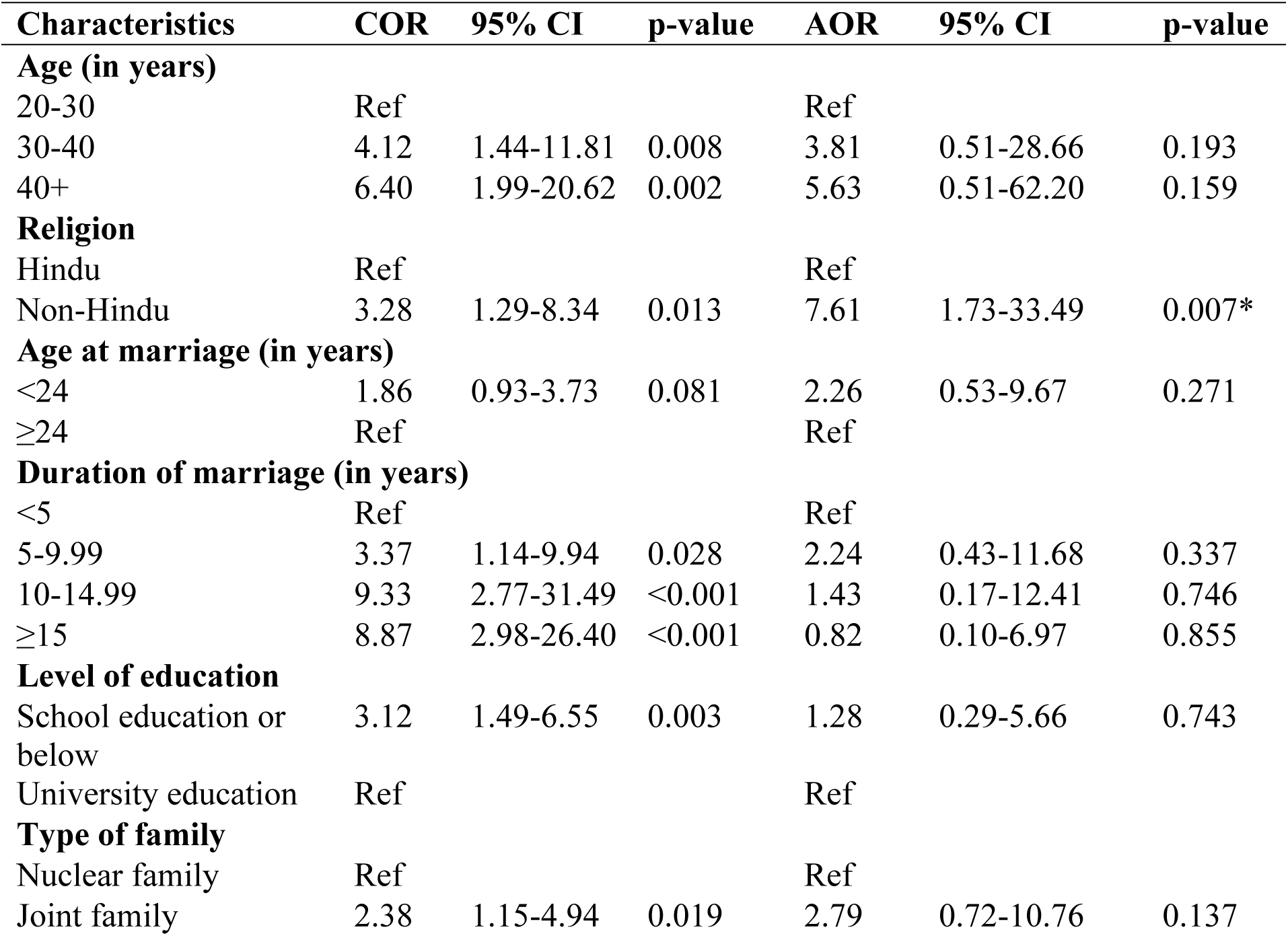
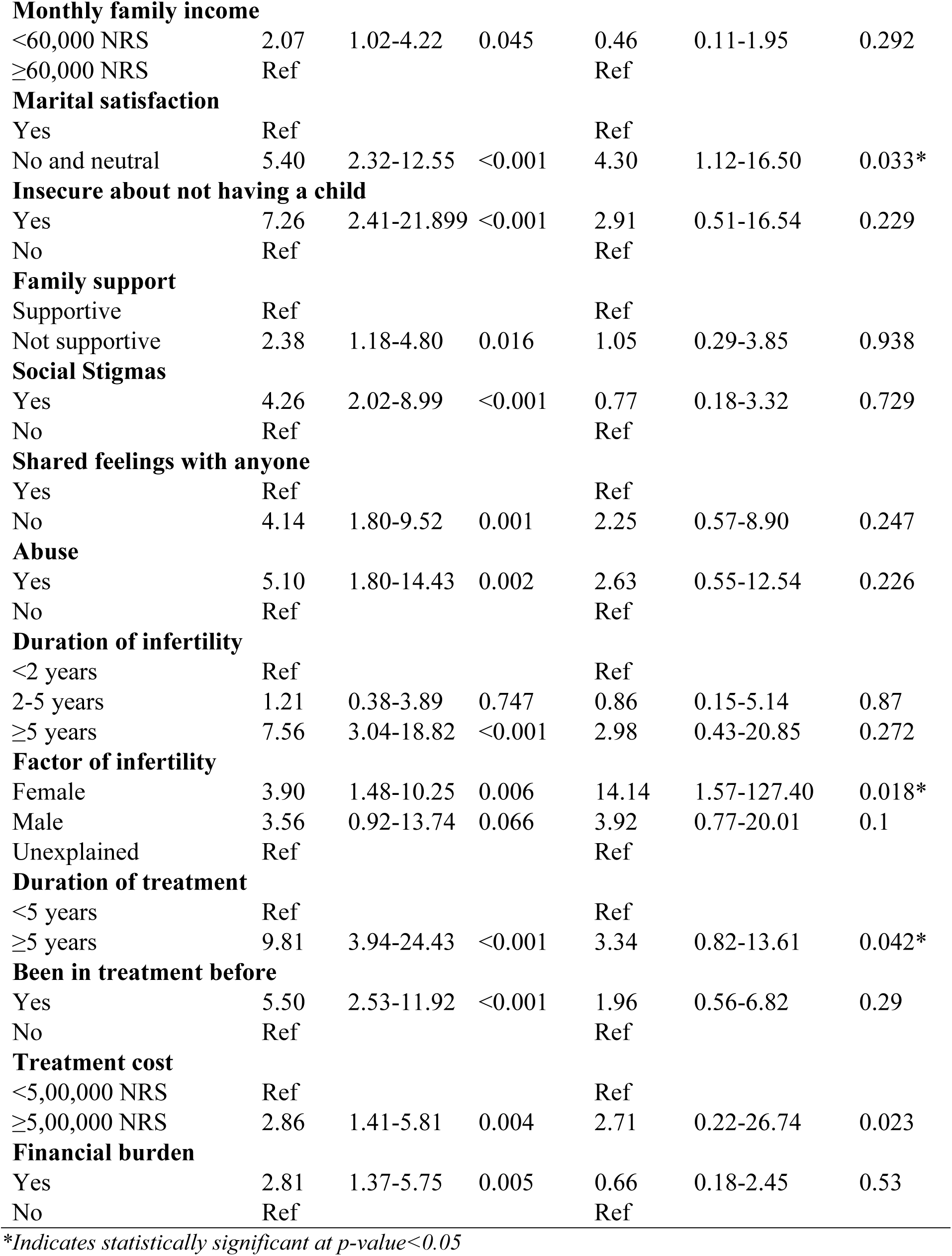
Multiple Logistics Regression between Depression and associated factors.

| Characteristics | COR | 95% CI | p-value | AOR | 95% CI | p-value |
| --- | --- | --- | --- | --- | --- | --- |
| **Age (in years)** |  |  |  |  |  |  |
| 20-30 | Ref |  |  | Ref |  |  |
| 30-40 | 4.12 | 1.44-11.81 | 0.008 | 3.81 | 0.51-28.66 | 0.193 |
| 40+ | 6.40 | 1.99-20.62 | 0.002 | 5.63 | 0.51-62.20 | 0.159 |
| **Religion** |  |  |  |  |  |  |
| Hindu | Ref |  |  | Ref |  |  |
| Non-Hindu | 3.28 | 1.29-8.34 | 0.013 | 7.61 | 1.73-33.49 | 0.007* |
| **Age at marriage (in years)** |  |  |  |  |  |  |
| <24 | 1.86 | 0.93-3.73 | 0.081 | 2.26 | 0.53-9.67 | 0.271 |
| ≥24 | Ref |  |  | Ref |  |  |
| **Duration of marriage (in years)** |  |  |  |  |  |  |
| <5 | Ref |  |  | Ref |  |  |
| 5-9.99 | 3.37 | 1.14-9.94 | 0.028 | 2.24 | 0.43-11.68 | 0.337 |
| 10-14.99 | 9.33 | 2.77-31.49 | <0.001 | 1.43 | 0.17-12.41 | 0.746 |
| ≥15 | 8.87 | 2.98-26.40 | <0.001 | 0.82 | 0.10-6.97 | 0.855 |
| **Level of education** |  |  |  |  |  |  |
| School education or below | 3.12 | 1.49-6.55 | 0.003 | 1.28 | 0.29-5.66 | 0.743 |
| University education | Ref |  |  | Ref |  |  |
| **Type of family** |  |  |  |  |  |  |
| Nuclear family | Ref |  |  | Ref |  |  |
| Joint family | 2.38 | 1.15-4.94 | 0.019 | 2.79 | 0.72-10.76 | 0.137 |
| **Monthly family income** |  |  |  |  |  |  |
| <60,000 NRS | 2.07 | 1.02-4.22 | 0.045 | 0.46 | 0.11-1.95 | 0.292 |
| ≥60,000 NRS | Ref |  |  | Ref |  |  |
| **Marital satisfaction** |  |  |  |  |  |  |
| Yes | Ref |  |  | Ref |  |  |
| No and neutral | 5.40 | 2.32-12.55 | <0.001 | 4.30 | 1.12-16.50 | 0.033* |
| **Insecure about not having a child** |  |  |  |  |  |  |
| Yes | 7.26 | 2.41-21.899 | <0.001 | 2.91 | 0.51-16.54 | 0.229 |
| No | Ref |  |  | Ref |  |  |
| **Family support** |  |  |  |  |  |  |
| Supportive | Ref |  |  | Ref |  |  |
| Not supportive | 2.38 | 1.18-4.80 | 0.016 | 1.05 | 0.29-3.85 | 0.938 |
| **Social Stigmas** |  |  |  |  |  |  |
| Yes | 4.26 | 2.02-8.99 | <0.001 | 0.77 | 0.18-3.32 | 0.729 |
| No | Ref |  |  | Ref |  |  |
| **Shared feelings with anyone** |  |  |  |  |  |  |
| Yes | Ref |  |  | Ref |  |  |
| No | 4.14 | 1.80-9.52 | 0.001 | 2.25 | 0.57-8.90 | 0.247 |
| **Abuse** |  |  |  |  |  |  |
| Yes | 5.10 | 1.80-14.43 | 0.002 | 2.63 | 0.55-12.54 | 0.226 |
| No | Ref |  |  | Ref |  |  |
| **Duration of infertility** |  |  |  |  |  |  |
| <2 years | Ref |  |  | Ref |  |  |
| 2-5 years | 1.21 | 0.38-3.89 | 0.747 | 0.86 | 0.15-5.14 | 0.87 |
| ≥5 years | 7.56 | 3.04-18.82 | <0.001 | 2.98 | 0.43-20.85 | 0.272 |
| **Factor of infertility** |  |  |  |  |  |  |
| Female | 3.90 | 1.48-10.25 | 0.006 | 14.14 | 1.57-127.40 | 0.018* |
| Male | 3.56 | 0.92-13.74 | 0.066 | 3.92 | 0.77-20.01 | 0.1 |
| Unexplained | Ref |  |  | Ref |  |  |
| **Duration of treatment** |  |  |  |  |  |  |
| <5 years | Ref |  |  | Ref |  |  |
| ≥5 years | 9.81 | 3.94-24.43 | <0.001 | 3.34 | 0.82-13.61 | 0.042* |
| **Been in treatment before** |  |  |  |  |  |  |
| Yes | 5.50 | 2.53-11.92 | <0.001 | 1.96 | 0.56-6.82 | 0.29 |
| No | Ref |  |  | Ref |  |  |
| **Treatment cost** |  |  |  |  |  |  |
| <5,00,000 NRS | Ref |  |  | Ref |  |  |
| ≥5,00,000 NRS | 2.86 | 1.41-5.81 | 0.004 | 2.71 | 0.22-26.74 | 0.023 |
| **Financial burden** |  |  |  |  |  |  |
| Yes | 2.81 | 1.37-5.75 | 0.005 | 0.66 | 0.18-2.45 | 0.53 |
| No | Ref |  |  | Ref |  |  |

*\*Indicates statistically significant at p-value<0.05*

## Discussion

The results of the current study assessed the multifaceted mental health status and associated experiences of infertile Nepalese women. More than 8 out of 10 respondents (82.5%) had some form of anxiety, more than half (51.2%) had some form of stress, and 35.0% of women had some form of depression, as found in the present study. Similar findings were found in a study conducted in Tehran, Iran, where 40.8% had depression and 86.8% had anxiety [13]. The study in Iran was also conducted in similar clinical settings among infertile women getting treatment for a longer duration, which may have aided in similar results like in the present study.

The findings in this study showed depression was significantly associated with duration of treatment, where women having treatment for more than 5 years were 3.34 times (AOR=3.34, 95% CI=0.82-13.61) more likely to have depression than women having treatment for less than 5 years. Similar findings were seen in a study conducted in Iran where anxiety and depression were most common after 4–6 years of infertility treatment (p=0.004) and severe depression could be found in those who had infertility treatment for 7–9 years [13]. As, due to the intensive duration of treatment procedures and multiple in-vitro fertilization failures, depression could have been seen after many years of infertility treatment in both studies.

The highest percentage of anxiety (82.5%), stress (51.7%), and level of depression (35.0%), which is significantly related to primary infertility, was found in this study. In contrast to the study conducted among infertile women (20-45 years of age) in a tertiary hospital in Pakistan with the highest level of depression (58.2%), anxiety (57.3%), and stress (50.0%) [15]. The findings of this study suggest that the difference in the prevalence may be due to differences in the treatment procedures and costs, with financial management of treatment costs varies with the country contexts.

The findings in this study showed a significant relationship of level of anxiety with the primary infertile group of women, where women who have never conceived were 2.58 times (AOR=2.58, 95% CI=1.65-36.09) more likely to have more anxiety issues than those who may have conceived earlier, significantly related to the female infertility factors. Similar findings were found in a study of assessment of depression, anxiety and stress among infertile women in Pakistan, where a significant relationship between the level of depression, anxiety, and stress (DASS) and the primary infertile group of women (p=0.049) was found [13]. The study in Pakistan was also conducted in similar settings among infertile women getting treatment, and where motherhood is the sole way for women to advance in life in both of the countries, which may have added on greater anxiety in women with female infertility factor.

In present study, women who were insecure about not having a child were 3.19 times (AOR=3.19, 95% CI=0.982-10.395) more likely to have stress than women who were not insecure. Whereas, in the study of Assessment of depression, anxiety and stress among infertile women in Pakistan, emotional strain of infertile women from in-laws was also associated with DASS (p=0.001) [13]. The study contradicts the findings of this study as family and husband support in treatment may be less in Muslim communities in Pakistan than in Nepal due to existing stereotypes and the status of women in Muslim countries.

This study depicts the financial burden (AOR=4.78, 95% CI=1.05-21.89) and marital satisfaction as significant (AOR=4.30, 95% CI=1.12-16.50) with depression level which was similar to the findings from a study in Iran in which among infertile women, age, social and sexual concern, marital relationship stress and financial stress were significantly related to distress [15]. This suggests high IVF treatment costs and no insurance system coverage for IVF as well as marital dissatisfaction, may have increased insecurity among women for not being able to have a child even after a long duration of marriage.

This research manifests an overall poor psychological status among the respondents regarding depression, anxiety, and stress. Infertility often leads to feelings of inadequacy, wherein this study, women who were insecure of not having a child were 3.194 times (AOR=3.19, 95% CI=0.98-10.40) more likely to have stress than those who weren’t insecure. Similar findings were seen in research in Iran, where infertility often leads to feelings of inadequacy, isolation, and disrupted self-identity, which, in turn, can trigger a cascade of psychological responses [32]. This suggests infertility has created the feeling of inadequacy and insecurity, and jealousy among women all around the world, which has affected the psychological state of women battling with it. This needs appropriate counseling techniques and social awareness campaigns by the government and/or policymakers to reduce the burden of poor mental health among infertile women, thus ensuring a better quality of life.

This study identified women facing social stigmas were 1.55 times (AOR=1.55, 95% CI=0.389-6.165) more likely to have anxiety than women not facing the social stigmas. While in a similar study conducted in Nigeria, social stigma was associated with higher rates of anxiety and depression [33]. Experiencing stigmatizing behavior (such as humiliation, discrimination, and devaluation) was associated with a threefold higher rate of depression [13]. Chinese women with infertility reported discrimination, shame, and reproductive pressures associated with depression [34].

Similarly, this study identified women having female factors of infertility were 14.14 times (AOR=14.14, 95% CI=1.57-127.40) more likely to have depression than those with an unexplained factor of infertility. This contradicts the findings of a cross-sectional study that was conducted in Tehran, Iran, where participants who had both/unknown causes of infertility were more likely to have poor well-being than men and women who had male and female factor infertility, respectively (OR = 1.54, 95% CI: 0.99–2.40), although this difference was not statistically significant (P = 0.053) [35]. Experiencing the infertility factor on her own grounds in such a stereotypical society might be a context in this study conducted among infertile women in Nepal, where the context could have been a lot easier in Iran, where stereotypical thoughts might not be the reason for poor psychological well-being.

Women having treatment duration more than 5 years were 3.34 times (AOR=3.34, 95% CI=0.82-13.61) more likely to have depression than women having treatment duration less than this as identified in the current study. It denotes that with increasing failed IVF pregnancies, women undergoing treatment suffer with increasing depression. In the similar study conducted by an Iranian university with 300 infertile women in three infertility centers, the mean psychological well-being score (PWB) in women who underwent IVF more than once was lower than the subjects who underwent IVF only once. Also, the mean PWB score in women who had more than once failed IVF pregnancies was lower than in subjects who had not had pregnancies by IVF. Also, the results of a qualitative study showed infertile women who receive IVF treatment because of receiving more hard treatment, failure treatment or failed IVF pregnancies feel out of control over their lives, which in turn reduces PWB [36].

This study had some limitations. Firstly, the information was collected from only one fertility center, however the clients attending this center were from all over the country. Additionally, other fertility centers had very few clients, and it was difficult to get permission. Confidential information about treatment procedures was difficult to assess, like factors of infertility and donor procedures, which may have influenced in the association in multiple regression in this current study. But this study has covered all the factors that may have affected the psychological distress among women dealing with fertility issues. This can aid in the development of specific and prompt psychological interventions, which are now lacking in the national public healthcare routine, for women battling infertility in order to help them manage potential mental health problems while meeting their reproductive goals.

## Conclusion

The majority of the women had anxiety, while half of the women had stress and nearly one-third of the women had some level of depression, which is significantly related to primary infertility. The findings showed the higher association between the mental health status of women with fertility issues with mostly social and treatment-related factors like insecurity, social stigma, abuse, parity, duration of treatment, factor of infertility, treatment cost and financial burden. These social and treatment-related factors create a complex interplay that impacts the mental health status of women facing fertility issues. Recognizing and addressing these factors within fertility care settings is crucial for providing comprehensive support and improving the overall well-being of women navigating infertility challenges. The findings underscore the importance of providing holistic support and tailored interventions to address the mental health needs of women experiencing fertility challenges, thereby enhancing their overall psychological well-being within the context of fertility care.

## Data Availability

Data is attached as a supplementary file.

## Abbreviations

AOR: Adjusted Odds Ratio
CI: Confidence Interval
COR: Crude Odds Ratio
DASS: Depression, Anxiety, and Stress Scale
IQR: Inter Quartile Range
IVF: In Vitro Fertilization
NRS: Nepalese Rupees
PWB: Psychological Well-being
SPSS: Statistical Package for Social Sciences
VIF: Variance Inflation Factor
WHO: World Health Organization

## Ethics approval and consent to participate

All methods of this study were carried out under the Helsinki Declaration of ethical principles for medical research involving human subjects. Ethical approval was taken from the Institutional Review Committee (IRC) of Manmohan Memorial Institute of Health Sciences (NEHCO/IRC/080/048). Permission to conduct the study was obtained from selected fertility centers in the Kathmandu Valley. The objectives, methods, and anticipated benefits of the study program were communicated to the participants prior to conducting the research. Written informed consent was obtained from each respondent, without any pressure or inducement of any kind, to ensure the voluntary participation of the respondents. Respondents were ensured voluntary participation and also freedom to withdraw from research at any time as per their comfort. The confidentiality of participants was maintained at every step of data collection, analysis, interpretation, and data dissemination. The personal information of the respondents was not taken to ensure confidentiality throughout the study process.

## Acknowledgement

Immense gratitude is conveyed to Vatsalya Health Care and the team for granting the permission to conduct research along with appreciative support. Gratitude is expressed towards the women dealing with fertility issues visiting the clinic for treatment who participated in the research.

## Supporting information

**S1 Text**

**S1 Data**

